# Frequency and Management of Drug and Chemical Poisoning among Children Attending an Emergency Department in a Single Hospital in Saudi Arabia

**DOI:** 10.1101/2020.09.08.20190868

**Authors:** Mansour Tobaiqy, Bandar A. Asiri, Ahmed H. Sholan, Yahya A. Alzahrani, Ayed A. Alkatheeri, Ahmed M. Mahha, Shamsia Alzahrani

**Author notes:** **Corresponding Author Dr. Mansour Tobaiqy**, BSc, MSc, Clin Pharmacol, PhD, PgCert, Assistant Professor, Department of Pharmacology, College of Medicine, University of Jeddah, P. O. Box 45311 Jeddah 21512. **Authors’ Contributions** MT conceived the study and, together with BAA, AHS, YAA, AAA, AMM and SA designed the questionnaire and performed the study. The manuscript was written by all authors.

## Abstract

**Background:** Unintentional poisoning is one of the common medical emergencies in children that leads to morbidity and mortality. Medications and chemical agents play a major role in these adverse events resulting in social, economic, and health consequences.

**Aims of the study:** The study aimed to evaluate the frequency and management of poisoning among children attending the emergency room at East Jeddah Hospital, Jeddah city, Saudi Arabia.

**Methods:** This study was a retrospective chart review of all acute pediatric poisoning incidences in children (0-16 years of age) from October-21-2016 to March-03-2020 who were admitted to the emergency department of East Jeddah Hospital, Jeddah city, Saudi Arabia. Data was analyzed via SPSS software.

**Results:** A total of 69 incidences of acute poisoning were admitted to the emergency room at East Jeddah Hospital; males were 55.1 %. Most children were under 5 years of age (59.4%). Unintentional poisoning occurred among 56.5% of observed cases of which 52.2% occurred in children younger than 5 years; 7.20% (5) patients were 12 to 16 years of age and had deliberate self-poisoning. The association between type of poisoning and age groups was statistically significant (Chi-square = 28.5057, p = 0.0001). Most incidences occurred at home (92.8%). Medicines were the most common cause of poisoning (73.9%). An excessive dose of prescribed medicine poisoning accidents was reported in 10.1% cases. Analgesics such as paracetamol were the most documented medication associated with poisoning (39.1%) followed by anticonvulsants and other central nervous system acting medicines (18.8 %). The most common route of poisoning was oral administration (81.2%). One mortality case was documented due to poisoning.

**Conclusion:** A total of 69 incidences of acute poisoning in children occurred in a single hospital over 3 years. Most incidents were unintentional and occurred in children younger than 5 years of age. Medicines were the most common cause of poisoning. Analgesics such as paracetamol were the most common documented medicine associated with poisoning.

## Introduction

Acute pediatric poisoning remains a worldwide health issue that requires medical attention at hospital emergency department with consequences of morbidity and mortality (1). It has social, economic, and health implications especially in children under the age of five who accounted for the largest percentage of poisonings globally (2).

The outcomes of poisoning ranged from mild incidences to severe complication or death, and most pediatric poisoning occurred accidentally by ingestion (3, 4). According to the World Health Organization (WHO) report (2002), an estimated 193,460 fatalities were linked to unintentional or accidental poisoning (5). In 2015, the American Association of Poison Control Center (AAPCC) reported that more than 1.3 million children were exposed to poisoning substances, and 40% of whom were children less than 3 years (6).

In Saudi Arabia, a review of seven years of case notes of children admitted to a single military hospital in Eastern province identified 168 accidental pediatric poisoning out of 9951 pediatric admissions (1.7%). This was most common in children between 1 to 3 years (63%). Most poisoning cases were related to medicines and accounted for 108 cases (64.3%) and household materials (n = 60, 35.7%) (7).

Another retrospective study of all pediatric poisoning cases reported to the drug and poison information center in Saudi Arabia identified 735 children presented to Pediatric Emergency department with poisoning from January 2010 to December 2016. Children younger than two years of age (n = 459, 62%) were significantly affected by poisoning.

Drug overdose (n = 119, 92.2%) was the major reason for poisoning and analgesics were the most common (n=26, 20.4%) (8).

The most frequent source of poisoning varied from country to country depending on social, economic, cultural, and educational background (1, 9). In developed countries, the most toxic substances are medicines and household products such as cleaning agents; developing countries frequently see kerosene and pesticides (1, 9). The current literature about poisoning among children in Saudi Arabia is limited in terms of examination of poisoning agent and the medications used for management and clinical outcomes.

## Aims

This study aimed to determine the frequency and management of acute pediatric poisoning in East Jeddah Hospital in Jeddah city, Saudi Arabia. The objectives of the study were:

- To identify the type of ingested poison.
- To describe the outcomes of poisoning.
- The name of antidote given.
- To describe the outcome of treatment.

## Methods

This study is a retrospective medical chart review of all acute pediatric poisoning cases admitted to the emergency department of East Jeddah Hospital in Jeddah city, Saudi Arabia. All children 16 years or younger admitted to as a result of acute poisoning from October 2016 to March 2020 were included.

### Data collection and statistical analysis

The data was collected from patients’ electronic medical reports at the hospital using data collection sheet by two pediatric physicians and two pharmacists who work at the emergency department. The data collection sheet was comprised of the patient’s demographic profile such as age, gender, weight, nationality, patient medical history, vital signs, the place when poisoning occurred, the type of poisoning, the type of medicine, route of intoxication, symptoms of intoxication, the medical treatment, antidote given, and the outcome of medical management. Data analysis used SPSS (SPSS Inc., Cary, NC version 20.0) and comprised descriptive statistics.

### Ethical Approval

Management authorization was gained from the Saudi Arabia Ministry of Health (MOH) Reference Number (01190) and ethical approval from King Abdulaziz City for Science and Technology (KACST), KSA: (H-02-J-002).

## Results

There were 69 incidences of acute poisoning.

### Characteristics of study population

There were 38 boys (55.1%) and 31 girls (44.9%). Most were under 5 years of age (n = 41, 59.4%), and most were Saudi citizens (n = 61, 88.4; Table 1).

**Table 1:** Demographic profile of children with poisoning (n = 69)

| Profile | No of children | % of children |
| --- | --- | --- |
| **Gender** |  |  |
| Male | 38 | 55.07 |
| Female | 31 | 44.93 |
| **Age group** |  |  |
| 0-5yrs | 41 | 59.42 |
| 6-11yrs | 18 | 26.09 |
| 12-16yrs | 10 | 14.49 |
| **Nationality** |  |  |
| Saudi | 61 | 88.41 |
| Non-Saudi | 8 | 11.59 |
| Total | 69 | 100.00 |

### Patterns of poisoning

Nearly half of the documented poisoning cases occurred unintentionally (n = 39, 56.5%), and were for children younger than 5 years of age (n = 36, 52.2%). The vast majority of these incidents occurred at home (n = 64, 92.8%). Intentional poisoning occurred in five cases (7.2%) for children over 5 years of age. Acute poisoning due to excessive dose of a prescribed medicines was reported in (n = 7, 10.1%) of the incidents, and (n = 18, 26.1%) were reported as unclassified (Table 2). The association between the pattern of poisoning and age groups was statistically significant (Chi-square = 28.5057, p = 0.0001) at 5% level of significance (Table 2, Figure 1)

**Table 2:** Patterns of poisoning incidences according to causes of toxic exposures in relation to age (n = 69)

| Causes / Age | (0-5) Year<br>n (%) | (6-11) Year<br>n (%) | (12-16) Year<br>n (%) | Total<br>n (%) |
| --- | --- | --- | --- | --- |
| Unintentional Poisoning | 36 (52.2) | 3 (4.3) | 0 (0) | 39 (56.5) |
| Deliberate Self-Poisoning | 0 (0) | 0 (0) | 5 (7.2) | 5 (7.2) |
| Excessive dose of a prescribed medicine | 3 (4.3) | 4 (5.8) | 0 (0) | 7 (10.1) |
| Unclassified | 3 (4.3) | 11 (15.9) | 4 (5.8) | 18 (26.1) |
| Chi-square=28.5057, p=0.0001* |  |  |  |  |

\*p<0.05

**Figure 1:**
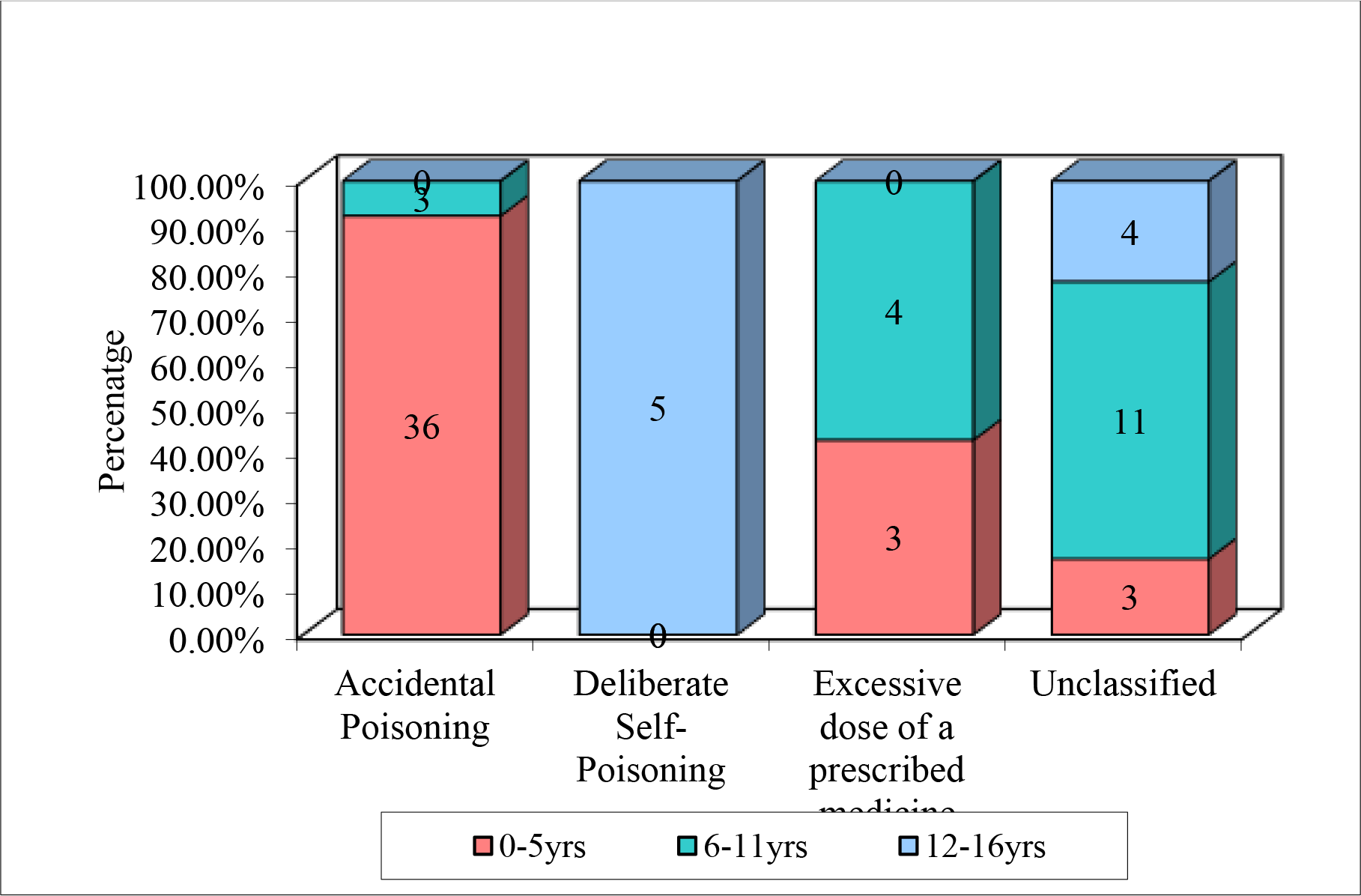
Patterns of poisoning incidences according to causes of toxic exposures in relation to age

### Sources and routs of poisoning

The main source of acute poisoning were medicines that were associated with the majority of incidents (n = 51, 73.9%) followed by intoxication by chemical substances (n = 9, 13%); nine cases (13%) were unknown toxic materials. Oral route of ingestion was found to be the most common route of intoxication in (n = 56, 81.2%) of all incidents as shown (Table 3; Figure 2).

**Table 3:** Summary of the site, types of poisons and route of intoxication (n = 69)

|  | No of children | % of children |
| --- | --- | --- |
| **Site of Poisoning** |  |  |
| At Home | 64 | 92.75 |
| At School | 1 | 1.45 |
| At Hospital | 4 | 5.80 |
| **Type of poisoning** |  |  |
| Medicines | 51 | 73.91 |
| Chemicals | 9 | 13.04 |
| Other | 9 | 13.04 |
| **Route of Intoxication** |  |  |
| Oral | 56 | 81.16 |
| Local | 4 | 5.80 |
| IV | 2 | 2.90 |
| Inhalation | 1 | 1.45 |
| Unknown | 6 | 8.70 |
| Total | 69 | 100.00 |

- Out of total of 69 patients 92.5% (64) patients had poisoning at home, 5.80% (4) at hospital followed by only 1 at school. - 73.91% (51) got poisoning by medicines followed by 13.04% (9). - 81.16% (56) patients had Oral route of Intoxication followed by local, IV, and inhalation.

**Figure 2:**
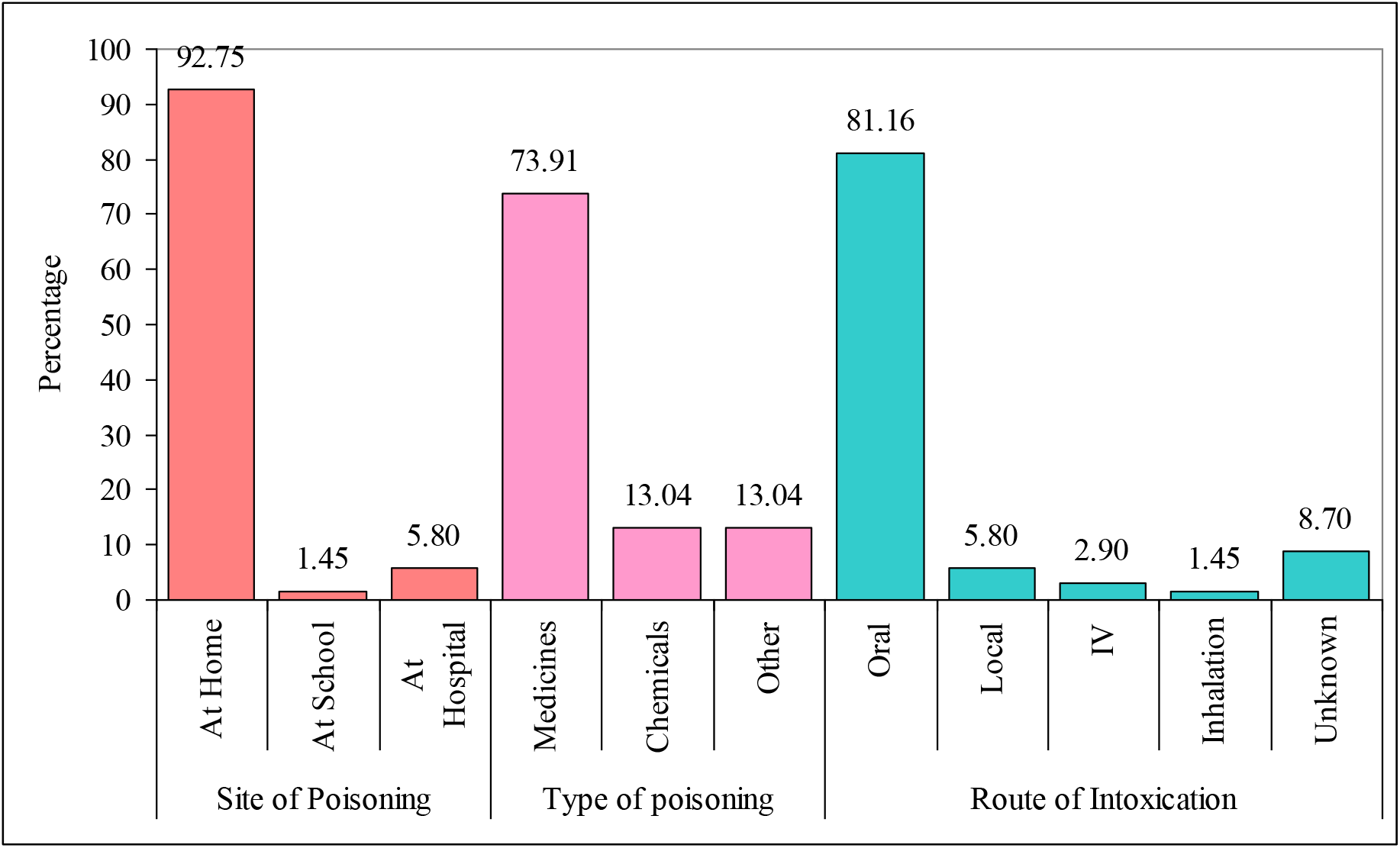
Distribution of children by site, types of poisons and route of intoxication

Analgesics and antipyretics such as paracetamol were the most common pharmaceutical agents that caused poisoning (n = 27, 39.1%) followed by central nervous system (CNS) active medicines such as anticonvulsants/CNS acting medicines (n = 13, 18.8%), antipsychotics (n = 9, 13%), cardiovascular medicines (n = 3, 4.3%), and one incidence of poisoning by antihistamines (n = 1, 1.4%). Chemical poisoning (n = 9, 13%) was caused by heavy metals and organophosphate compounds. Seven incidents of chemical poisoning (10.1%) occurred in which the exact type of chemical had not been identified as shown (Table 4).

**Table 4:** Agents involved in toxic exposures (n=69)

|  | No of children | % of children |
| --- | --- | --- |
| **Class of medicine** |  |  |
| Analgesics / Antipyretics | 27 | 39.10 |
| Anticonvulsants/CNS Drugs | 13 | 18.80 |
| Antipsychotics | 9 | 13.00 |
| Antihistamines | 1 | 1.40 |
| **Chemical substances** |  |  |
| Heavy Metals | 3 | 4.30 |
| Organic and hydrocarbon compound | 3 | 4.30 |
| Organophosphate | 2 | 2.90 |
| Alcohol/ Methanol | 1 | 1.40 |
| Unknown | 7 | 10.10 |

### Management of poisoning

Treatment of poisoning incidents varied from one case to another depending on patient condition, type of poisoning and time of exposure. In this study, treatment intervention was reported in most cases as shown in (Table 5). Antidote pharmacological treatment was given to 31 patients (44.9%), supportive treatment in 15 (21.7%), however 23 (33.3%) required no medical intervention. Of those who received antidotes, (n = 16, 51.6%) were given activated charcoal, N-acetylcysteine (n = 9, 29%), antihistamines (n = 3, 9.7%). One patient received naloxone, atropine and fomepizole (Table 6).

**Table 5:** Treatment intervention (n = 69)

| Intervention | No of children | % of children |
| --- | --- | --- |
| Antidote | 31 | 44.90 |
| Supportive Treatment | 15 | 21.70 |
| No need | 23 | 33.30 |
| Total | 69 | 100.00 |

**Table 6:** Antidote involved in cases treatment (n = 31)

| Antidote | No of children | % of children |
| --- | --- | --- |
| Activated Charcoal | 16 | 51.60 |
| N-Acetylcysteine | 9 | 29.00 |
| Antihistamine | 3 | 9.70 |
| Naloxone | 1 | 3.20 |
| Atropine | 1 | 3.20 |
| Fomepizole | 1 | 3.20 |

### Outcome of poisoning

A total of 25 (36.2%) children were admitted to the pediatric ward, while 35 (50.7%) were discharged from the emergency room after receiving treatment and eight (11.6%) were admitted to the Pediatric Intensive Care Unit (PICU). One case of mortality was documented (n = 1, 1.4%) (Table 7; Figure 3).

**Table 7:** Disposition and outcomes of toxic exposures cases (n = 69)

| Disposition | No of children | % of children |
| --- | --- | --- |
| Discharged | 35 | 50.7 |
| Paediatric wards | 25 | 36.2 |
| Paediatric Intensive Care Unit (PICU) | 8 | 11.6 |
| Died | 1 | 1.4 |

**Figure 3:**
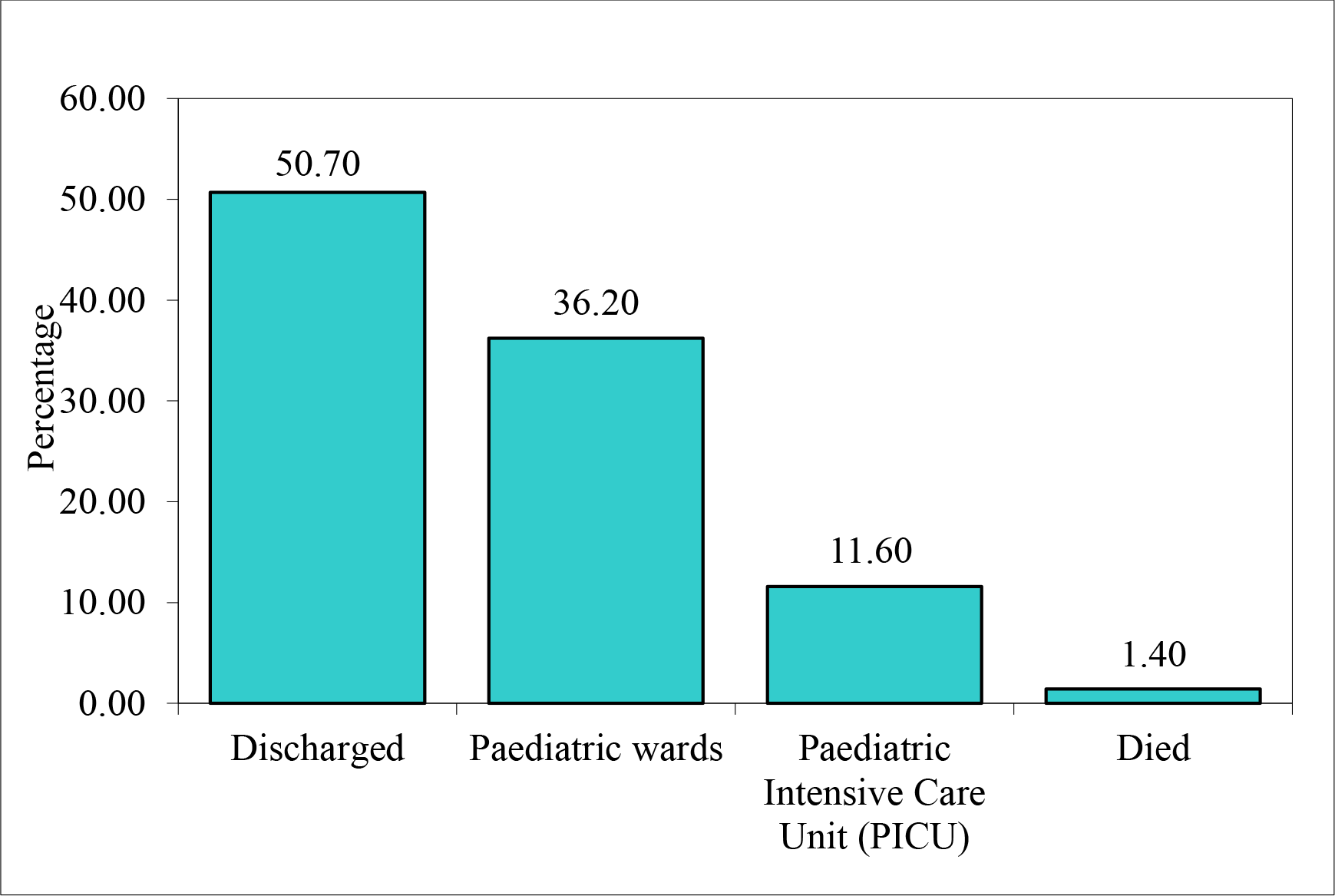
Disposition and outcomes of toxic exposures cases

## Discussion

There were 69 incidences of acute poisoning admitted to the emergency room at a single Hospital in Jeddah, Saudi Arabia over ∼3.5 years. Most incidents were in boys consistent with a previous study (10). Although there have been no comprehensive epidemiological studies of the incidence of poisoning in the Kingdom of Saudi Arabia to date, studies published for different regions and time periods also indicate that children younger than 5 years are most often affected; similar result were shown here (11).

In the literature, most incidences of poisoning occurred at home, and males had a greater risk for poisoning. Unintentional poisoning accounted for most incidents in this study (56.5%) while intentional self-poisoning was reported in five incidents. Of note, attempted suicide is a criminal act in Saudi Arabia (12); social stigmatization associated with psychiatric illness and their medications is another issue (13), both may play a role in under-reporting of attempted intentional poisoning.

The vast majority of incidences occurred at home (92.8%). Unintentional poisoning occurred for several reasons including child’s curiosity, natural tendency to explore the environment, and a lack of awareness of surrounding risk. Guardians must be responsible for the selection of medicines and safely storing household supplies and educating young children about potential risk (14, 15). Intentional poisoning usually affected adolescent, in a study of 148 cases of poisoning, 86% were accidental; of the intentional cases, 33% were suicidal in subjects 12 years or older (14, 15).

In this study, ingested medicines were found to be the main cause of acute poisoning (73.9%) consistent with a previous study (16) followed by chemical poisoning that occurred in 13% of the incidents. According to the Annual Report of AAPCC (2018) National Poison Data System (NPDS) (6), the top five most common exposures in children age 5 years or less were cosmetics and personal care products (12.1%) followed by household cleaning substances (10.7%) and analgesics (9.04).

Analgesics—specifically non-steroidal anti-inflammatory drugs—as well as household cleaning substances are the most common poisons in children in recent studies. (17,18). In these studies, analgesics like paracetamol were the most documented cause of poisoning in children (39.1%) in addition to pharmaceutical products such as syrups that attract children due to colors and flavors (19) followed by anticonvulsants (18.8%) and antipsychotics (13%). Other studies reported that neurologic medicines were the most common drugs causing poisoning in children followed by analgesics (1).

The management of poisoning cases in pediatric cases depends on several factors such as the type of poison, the dose, clinical manifestation, age, presence of other diseases or injury, and the time of poisoning exposure (20).

Most poisoning cases were managed in the hospital through decontamination with active charcoal (51.6%), and N-acetylcysteine (antidote of paracetamol) was given to (29%) of the cases. Previous studies have shown that active charcoal alone is a better treatment for poisoning cases presenting in the hospital within one hour (21). Activated charcoal can decrease the absorption in stomach and intestine for a wide variety of toxins and medicines such as carbamazepine, phenobarbital, theophylline, salicylates, and valproic acid (20),

In this study, 25 patients (36.2%) were admitted to the pediatric ward while (n = 8, 11.6%) cases were admitted to the PICU; one mortality was documented. The clinical severity of poisoning incidents in this study is higher than what is reported in similar research where most cases where mild, and 101 (17.2%) cases were admitted to the hospital and only 21 (3.6%) were admitted to the PICU (1).

Of note, most childhood poisoning incidences occurred due to the easy availability of medicines and chemicals at home in several forms and a lack of parental supervision to keep these materials in a safe place and out of reach to children. A World Health Organization report revealed that an estimated 193,460 deaths were caused annually because of unintentional poisoning worldwide of which 84% occurred in low- and middle-income countries. Educating the community and particularly parents about the risks of drugs and chemicals poisoning in children may reduce the occurrence of such harmful adverse events (16, 22, 23).

One limitation of this study is its retrospective nature and single setting. Incomplete and missing data may also limit the generalizability of the study’s findings. The results highlight the importance of community education and developing guidelines for monitoring and managing poisoning incidents in Saudi Arabia.

## Conclusions

There were 69 incidences of acute poisoning in children in a single hospital within approximately three years. Most incidents were unintentional and occurred in children younger than 5 years of age. Most poisonings were due to medicines. Analgesics such as paracetamol was the most common documented medicine associated with poisoning. Activated charcoal was the most common treatment. Educating the community and particularly parents about the risks of drugs and chemicals poisoning in children, may reduce the occurrence of such harmful adverse events

## Data Availability

Data are available from the corresponding author on reasonable request.

From the results of the above table, 52.20% (36) patients 0-5yrs age group had accidental poisoning, 7.20% (5) patients 12-16yrs of age group had deliberate self-poisoning followed by unclassified causes in most of the patients in all the age groups. The association between the pattern of poisoning and age groups was found to be statistically significant (Chi-square = 28.5057, p = 0.0001) at 5% level of significance.

- Out of total of 69 patients 92.5% (64) patients had poisoning at home, 5.80% (4) at hospital followed by only 1 at school.
- 73.91% (51) got poisoning by medicines followed by 13.04% (9).
- 81.16% (56) patients had Oral route of Intoxication followed by local, IV, and inhalation.

